# Perceptions and Insights: A Qualitative Assessment of an AI-Assisted Psychiatric Triage System Implemented in an Outpatient Hospital Setting

**DOI:** 10.1101/2025.03.04.25323387

**Authors:** Oleksandr Knyahnytskyi, Jazmin Eadie, Kimia Asadpour, Callum Stephenson, Megan Yang, Taras Reshetukha, Christina Moi, Tricia Barrett, Meghanne Hicks, Gilmar Gutierrez, Anchan Kumar, Jasleen Jagayat, Saad Sajid, Charmy Patel, Christina Holmes, Amir Shirazi, Vedat Verter, Claudio Soares, Mohsen Omrani, Nazanin Alavi

**Affiliations:** Department of Psychiatry, Faculty of Health Sciences, Queen’s University, Kingston, Canada; Kingston Health Sciences Centre, Kingston ON, Canada; Centre for Neuroscience Studies, Faculty of Health Sciences, Queen’s University, Kingston ON, Canada; OPTT Health, Kingston ON, Canada; Smith School of Business, Queen’s University, Kingston ON, Canada

**Keywords:** Online cognitive behavioural therapy, Mental Health, Artificial Intelligence, Psychiatry, Psychotherapy, Cognitive behavioural therapy, Qualitative, Perceptions and Experiences

## Abstract

**Introduction:** The Canadian healthcare system is approaching a breaking point. With mental health being a leading cause of disability, innovative solutions are necessary to provide adequate care. Digital mental health programs, such as electronic cognitive behavioural therapy (eCBT), have proven effective in reducing the challenges of traditional psychotherapy such as long waitlists, stigma, geographic barriers, and reducing time constraints. Furthermore, artificial intelligence (AI) has shown potential utility within the healthcare system, particularly in treatment recommendations and improving patient engagement. Despite the benefits that AI and digital mental health programs provide, they are rarely implemented in real-world healthcare settings.

**Objective:** This study aims to explore patient experiences and perceptions of an AI-assisted triage system paired with a digital psychotherapy program. The objective is to highlight the potential modifiable barriers to implementing these digital systems in real-world healthcare settings.

**Methods:** 45 adult outpatient psychiatry patients (n=45) who used an AI-assisted triaging system and digital psychotherapy modules, were surveyed through Qualtrics. This survey examined their perceptions of AI within mental healthcare, its utility within triaging, and their experiences with the digital psychotherapy program. Free-text survey responses were independently coded and analyzed using thematic analysis.

**Results:** Thematic analysis revealed three major themes for client’s perceptions of the AI-assisted triage system: (1) AI as a replacement, (2) the utility of AI, and (3) AI complexity recognition. For the digital psychotherapy program, the themes were: (1) interactions with technology, (2) online therapy program structure, and (3) differential user experience.

**Conclusion:** Participants highlighted the importance of human oversight to ensure accuracy and liked that the AI allowed them to access care faster. Suggestions for improving the digital psychotherapy program included enhancing user-friendliness, increasing human contact, and making it more accessible for neurodivergence.

## Introduction

### Background Information

The current state of primary healthcare in Canada, especially post-COVID-19 pandemic, has been slowly approaching a breaking point with many considering it to be in crisis.^1^ Despite general improvements in the quality of care over the years, timely access to care, efficient services, and consistent care have been brought up as areas of concern in the current system.^2,3^ With mental illness being a leading cause of disability in Canada, these concerns are further amplified due to a distinct lack of mental health resources in both clinical and public settings.^4^ Roughly 6.7 million Canadians are affected by mental health problems annually, with around half reporting unmet or partially unmet healthcare needs.^5-7^ The lack of readily available resources has resulted in a severe socioeconomic burden that calls for innovative solutions.^2,4^

Digital mental health interventions aim to address these issues by making healthcare services more accessible, helping streamline the workflow of hospitals, and access to care.^8^ Interventions such as online cognitive behavioural therapy (eCBT), have emerged as effective solutions to address gaps in mental health care accessibility.^9^ eCBT involves structured, evidence-based therapy modules delivered through online platforms, allowing individuals to engage in cognitive and behavioural strategies to manage mental health challenges.^10^ eCBT specifically addresses concerns of accessibility, as it helps overcome barriers such as geographic limitations, long wait times, and stigma associated with seeking in-person care^11^. Additionally, eCBT minimizes the time needed for direct clinician involvement, which in turn helps reduce the financial burden on the patients, and alleviates pressure on overburdened mental health services, helping address systemic inefficiencies.^10,12^ With the digital nature of interventions such as eCBT, it becomes an ideal candidate for integrating more novel and advanced technologies, such as artificial intelligence (AI), to help further optimize care pathways.

In recent years, AI has seen a notable increase in utility within healthcare settings due to its effectiveness in tasks such as treatment recommendations, patient engagement and adherence, and therapy effectiveness.^8,13^ Artificial intelligence is a scientific domain that is primarily focused on the creation of “intelligent” machines through the development of algorithms or sets of rules.^14^ One of the strengths of AI, relating to healthcare, is its ability to identify and learn various complex patterns through the analysis of multimodal and multidimensional datasets.^14^ More specifically, branches of AI, such as Machine Learning (ML) or Natural Language Processing (NLP), are often utilized within healthcare settings due to their ability to recognize dialectical patterns. In turn, this recognition helps inform the development of treatment protocols, illness classification, and other precise medical applications.^13^ These AI functionalities have been the focus of the Queen’s University Online Psychotherapy Lab (QUOPL), which established and implemented an innovative digital mental health intervention using a joint AI-assisted triage and online psychotherapy program at Kingston Health Sciences Centre (KHSC). This system uses NLP models to analyze patient narratives related to their mental health challenges, suggesting the most appropriate level of care and treatment program.^15^ Through a series of retrospective and ongoing clinical trials, the implementation of this algorithm has been shown to effectively assist in clinical decision-making, optimize resource utilization, reduce psychiatrist wait times, and provide timely access to effective online mental health services tailored to the patient’s symptoms and care needs.^16^

To further assess the effectiveness and implementation of this algorithm, a quality improvement (QI) study, funded by the Ontario Centre of Innovation (OCI), was conducted from October 2023 to March 2024. This study, *“Using AI-Assisted Triage & Online Psychotherapy to Address Access to Mental Health Care,”* occurred at the mental health ambulatory services of KHSC, Hotel Dieu Hospital (HDH) site. That study aimed to evaluate the efficiency and feasibility of the AI-assisted triage algorithm in managing referrals from primary care providers. Specifically, the study examined the proportion of patients requiring urgent psychiatric consultation post-referral and assessed the appropriateness of wait times to see a psychiatrist across different care levels (Figure 1).

**Figure 1.**
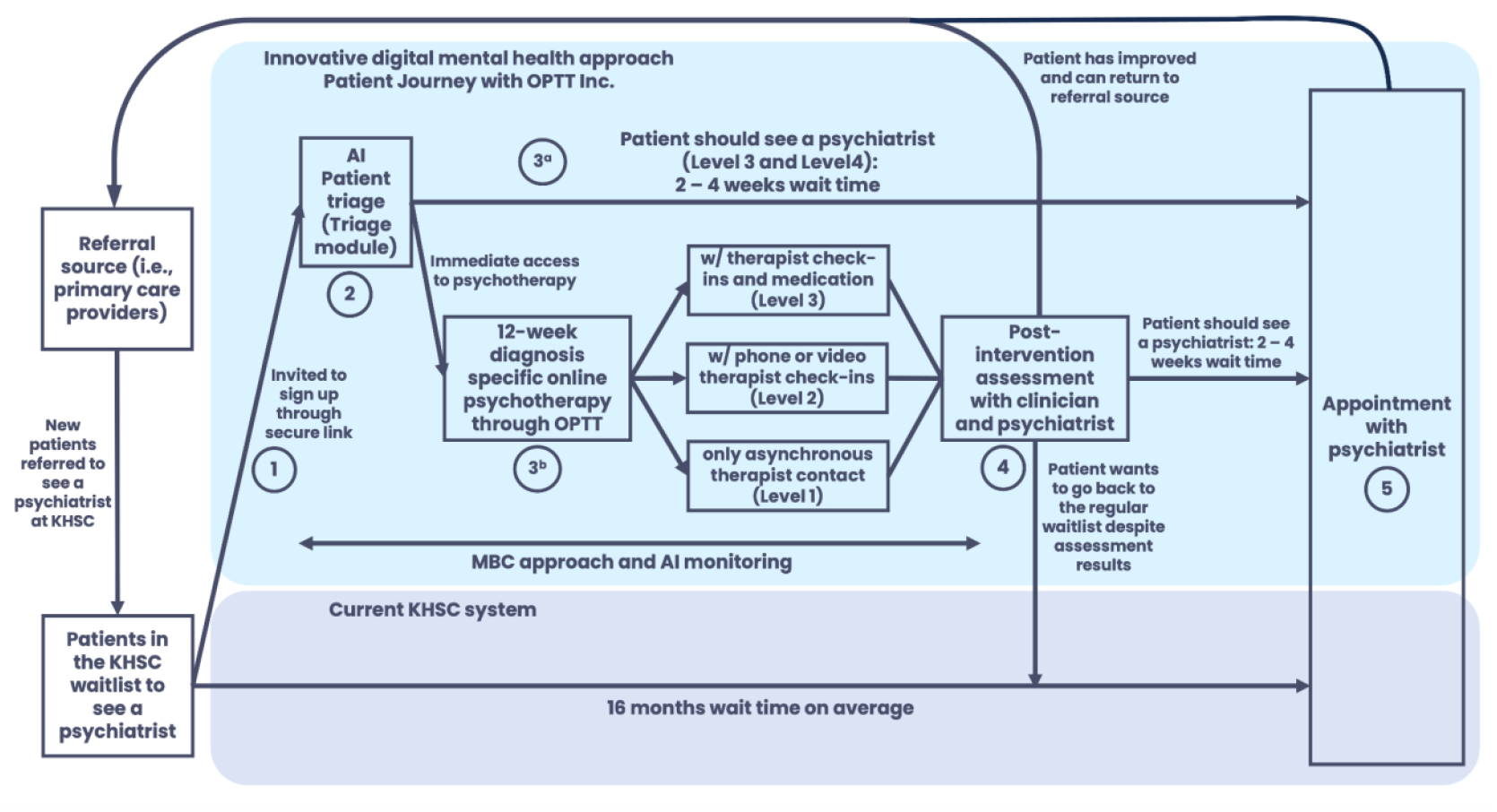
*Note*. Patient journey as part of the QI study conducted by our team at QUOPL. Figure outlines (1) the patient journey in the current triage system at KHSC and (2) the patient journey through the AI-triage approach.

In the current study, we conducted a qualitative exploration of the personal experiences of individuals who engaged with the AI-assisted triage system and subsequent digital psychotherapy programs. The use of qualitative data in healthcare research is important as it focuses on knowledge grounded in human experience, and allows for a better understanding of the social, and cultural dimensions associated with healthcare.^17^ Qualitative methods serve as a key tool in implementation research since they help explain complex phenomena surrounding user experience and perceptions.^18^ As a result, qualitative methods can inform future implementations of certain products, protocols, or systems.^18^

In this study we sought to understand patient experiences with the AI-assisted triage system and the digital psychotherapy program. Using qualitative methodology, we aimed to explain complex phenomena surrounding user experience and perceptions to understand the barriers associated with the implementation and acceptability of interventions such as AI triaging systems and digital psychotherapy programs. Through addressing the barriers to implementation, we hope to improve future applications of these interventions.

## Materials & Methods

### Participants and study design

A total of 101 patients participated in our QI study implemented at KHSC from October 2023 to March 2024. Participants were adult patients (≧18 years of age), who had been referred by family physicians to the ambulatory clinic at KHSC, HDH in Kingston ON. Patients on the waitlist, referred between January and September 2023, were invited to participate in the study utilizing an AI-assisted triage tool paired with digital psychotherapy modules. Upon completion or drop out from the initial study, participants were offered the option to complete an anonymous Qualtrics survey outlining their thoughts and experiences with both the AI-assisted triage and the digital psychotherapy program. Of the initial 101 individuals who participated in the QI study, 45 completed the Qualtrics Survey.

The AI-assisted triage tool and digital psychotherapy module platform, designed by MO, NA, and the research team, was hosted on the secured cloud-based platform OPTT.^19^ Participants received a link to the triage module, which featured narratives about mental health disorders through the stories of characters, illustrating how these characters navigated their journeys and shared their experiences. Patients were then encouraged to share their own stories and completed a series of questionnaires.

The triage tool assessed each patient’s needs, suggesting a level of care (mild, moderate, or severe) and determined the urgency and necessity of seeing a psychiatrist. It also recommended suitable digital psychotherapy options, and the level of support required in psychotherapy, such as fully online sessions, online therapy with weekly therapist check-ins, or online therapy with weekly check-ins and psychiatrist appointments. All AI-assisted assessments were finalized by a practicing psychiatrist to ensure accuracy.

### Intervention

For this study, 45 adult participants (n=45, 18 years old and above) directly involved in the use of the AI-triage system, received an anonymous Qualtrics survey link through an email address they had previously provided. Before beginning the questionnaire, the survey’s purpose and procedures were clearly explained, and participants reconfirmed their consent to proceed. Ethical approval for this study was obtained from the Queen’s University Health Sciences and Affiliated Teaching Hospitals Research Ethics Board in Kingston, ON, Canada.^20^

### Survey Development

The post-participation survey was designed based on the framework of previous surveys implemented by the research team to capture the user experiences of participants taking part in AI-assisted triage and the digital psychotherapy program.^21^ The content was created through comprehensive literature reviews and validated qualitative assessment tools. Before distribution to the participants, the survey underwent several rounds of revision by team members with expertise in AI and online psychotherapy.

### Data Analysis

The post-participation survey was created and analyzed using Qualtrics.^22^ The primary method of analysis for the open-text responses within the survey utilized a phenomenological approach. More specifically, thematic analysis was used to analyze survey responses. Thematic analysis is a common qualitative methodology used to identify and interpret major patterns within a text-based data set.^23^ The use of thematic analysis allows for the emergence of themes or “patterned responses” from within a data set, which in turn can help guide our understanding of complex phenomena such as patient perceptions and experiences.^24^ Qualitative data analysis began with the compilation of all the participants’ responses into a single spreadsheet. A complete readthrough of the participant responses was then independently conducted by trained research personnel to get a general sense of the participants’ perspectives and experiences. Research personnel then independently coded the open-text data via open coding, creating a preliminary list of codes and potential themes. A standardized code was then collaboratively developed through a thorough review of each independently developed coding scheme. Discrepancies were resolved through discussion between coders, to create a final standardized code. Using the standardized code, the remaining entries were coded to extract the final themes and subthemes within the data. Key themes were created through the organization of emergent primary, secondary, and tertiary codes obtained from the data.

## Results

Data extracted from the Qualtrics survey that examined patient perceptions of the AI-assisted triage and eCBT program revealed six themes. Three themes related to the use of an AI-assisted triage system: 1) AI as a replacement, 2) Utility of AI, and 3) AI Complexity Recognition emerged. Additionally, there were three themes related to the eCBT program: 1) Interactions with Technology, 2) Online Therapy Program Structure, and 3) Differential User Experience. The themes and their associated subthemes are described in more detail below.

### Participant Experiences and Perceptions of the AI-assisted Triage

Theme: AI as a Replacement

Subtheme: Need for and Potential of AI in Healthcare

Participants expressed varied opinions regarding the implementation of AI-assisted systems within real-world healthcare settings. Many participants supported the use of such a system, focusing on its utility as a gateway towards accessible care: *“Whatever tool helps connect mental health professionals with patients in need is all goodness.”* Another participant shared a similar sentiment, emphasizing the necessity of AI due to deficiencies in our current healthcare systems: *“It’s [AI] necessary at this point since there are not enough care providers.”* On the other hand, some participants focused on the potential limitations of AI and its use in a mental healthcare setting: *“AI is improving, but humans are complicated therefore AI is invalid to treat real people.”*

Subtheme: Need for Human Oversight

Relating to the need for, and potential of AI, participants highlighted a need for human oversight in the implementation of an AI-assisted healthcare system. One patient mentioned the importance of human oversight in monitoring the behaviour of the system:

> *“I work in AI, and I think that it can be very beneficial in health care, but it is crucial to have a human in the loop and to monitor the behaviour of the systems consistently.”*

Additionally, other participants discussed the necessity of human control over diagnosis and medication prescription: “*It [AI] has potential, but there has to be access to somebody who can diagnose and prescribe medications.”* When discussing the differences between traditional and AI-assisted triage, one participant mentioned that the AI-assisted triage system was *“Ok -but it can miss that ‘gut’ feeling & anyone or anything that fits outside the box.”*

Theme: Applicability of AI

Subtheme: Utility

Participants generally supported the use of AI to improve the efficiency of the care provided in a healthcare setting. *“I believe efficiency in the service of fast-tracking patients to different forms of care is important and if AI helps with that, then I have no problem with it.”* When discussing the differences between the care the participants have previously received and the AI-assisted system, one participant mentioned “*I could not access a psychiatrist through my family doctor, so this intake trial was an enabler for me.”*. Additionally, another participant mentioned that one of the benefits of the system was that *“I got the help and medication I needed after 37 years.”* In contrast, some participants were not as open to the application of AI highlighting a limited acceptability of its utility: *“For intake, I would accept AI, but for diagnosis or ongoing treatment, I would NOT support AI at this time.”*

Subtheme: Connection

When it came to discussing the key limitations of the AI-assisted triage system, some participants discussed a lack of personal connection as being a key constraint in their acceptability of the use of AI: *“I do not feel anyone can get a truly deep connection with an AI facilitated healthcare program - it was not for me.”* However, participants mentioned this in a more general sense regarding their views of AI in healthcare as a whole: *“When care and social interactions are compromised, I disagree with having AI in healthcare. Though, when it is used positively in [this] way, I felt it was used in this program*.

Theme: AI Complexity Recognition

Subtheme: Unaddressed Needs

In general, participants felt that AI is not able to account for the complexity of mental illness to the same degree as a human. As a result, this might have led to unaddressed needs in some of the participants. One participant mentioned *“The program diagnosed me as depressed, but my depression stems from untreated ADHD. It was explained to me that there is an ADHD module that maybe was not ready yet.”* Another participant also highlighted an issue in their unmet needs, though with a specific focus on the speed and utility of the AI system: *“I am not sure the speed is relevant. I just do not see how a quicker service would be of use if the service is not addressing my needs.”*

Subtheme: AI’s Limitations

Though participants were generally open to AI, many discussed various limitations to its use, highlighting human complexity, the lack of specialized programs, and unmet needs as reasons that AI was not a favourable alternative.

> *“I am concerned for others that the AI diagnosed me with depression when my real problem is ADHD.”*
>
> *“Okay-but it can miss that “gut” feeling & anyone or anything that fits outside the box.”*
>
> *“AI is improving, but humans are complicated, therefore AI is invalid to treat real people.”*

### Participant experiences and perceptions of the digital psychotherapy program

Theme: Interactions with Technology

Subtheme: Problems with the technology

Participants expressed various frustrations and challenges regarding the user experience with the platform. Common issues included difficulties logging in, an inability to access or read feedback, receiving incorrect modules, and not having an *“intuitive user design.”*

> *“There are a lot of kinks in the system that need to be worked out to keep users from getting frustrated. I experienced login issues, and I found some of the User Experience challenging to understand at times.”*
>
> *“My biggest issue was with the technical problems. For someone like me, problems like not being able to read the feedback, or being given the wrong module are enough to derail the momentum and gave me a very convenient excuse to give up.”*

Theme: Online Therapy Program Structure

Subtheme: Convenience, Flexibility, Diversity and Accessibility

Clients expressed mixed feelings about the program’s level of convenience, flexibility, and accessibility. Some appreciated the ability to express themselves privately without the immediate presence of a care provider, commenting that this feature provided an outlet and access to care while they waited to see a psychiatrist. However, concerns were raised regarding the program’s ability to address neurodiversity, suggesting that neurodivergence needs might have gone unaddressed with the digital psychotherapy program.

> *“I like it, the fact that the professional is not there looking at you, and you can just write your heart out without having to worry about social cues is useful.”*
>
> *“I think it is great that people have an outlet while waiting for a psychiatrist”*
>
> *“Neurodivergence has gone unaddressed.”*

Subtheme: Program Repetitiveness and Strictness

Some participants found the content of the program to be repetitive. Others found the deadlines to be challenging, which resulted in them finding the program to be more of a hindrance resulting in various negative feelings such as anger.

> *“Repetitive info”*
>
> *“More of a hindrance than helpful.”*
>
> *“This program was frustrating. Between deadlines, difficulties navigating the online program & the repetitiveness, I do not recommend it. It made me more upset and angrier than I had before attempting the program in the first place.”*

Theme: Differential User Experience

Subtheme: Level of Helpfulness

Differing experiences occurred for participants in this program, with some being helped and others feeling like this program was a hindrance. Clients who found this program to be helpful enjoyed the feedback and found benefits in the ability to continually set goals. Some individuals commented on having received the care that they have been seeking for years.

> *“I got the help and medication that I needed after 37 years.”*
>
> *“Great doctors”*
>
> *“Feedback was helpful”*
>
> *“I found it beneficial to have set goals every week, and the feedback was reassuring. I found it challenging to remember all the content, there should be reminders in each session of the last session to reinforce the content.”*

In contrast, other participants felt like they were in the same place that they were in before starting the program. As a result of the therapeutic relationship, a participant expressed feeling like they needed to make stuff up to fit the homework.

> *“I had the hope that when I was done with the program, I would be connected to a psychiatrist right away, so I could continue the treatment. But now I am kind of back to where I was at the beginning. Having to go see a doctor at a walk-in clinic and hope that what they give me will work.”*
>
> *“… a few weeks into it, it became a task I needed to do to please the person calling me once a week. The experience was a bit like an educational course with due dates and assignments. I was completing the assignments to please the person calling me (like I was trying to get a good grade). I found myself making up things or thinking back into my past to come up with things that fit the assignments. I finally realized that completing assignments to please the person calling me is not what I should be doing to deal with and work through the issues I am trying to work through. This decision to quit the program has nothing to do with the person calling me. It is the program and how it is being presented.”*

Subtheme: Perceptions of the Need for Human Contact

Throughout the program, each participant was supported by a care provider who worked with them in various capacities. Everyone received feedback, some received weekly phone calls, and others got both of those and saw a psychiatrist. Participants had varying reactions to different levels of care. Some felt like the care provider kept them on track and appreciated how the weekly phone calls could clarify any confusing content from the eCBT. However, a common theme was that people who only received eCBT tended to feel like they missed out on the back-and-forth aspect of verbal communication.

> *“I appreciated that the [care provider] was there and kept me on track. If there had not been that human element I certainly would not have finished.”*
>
> *“All contact with people was great”*
>
> *“The benefits of having a knowledgeable mental health expert are unparalleled, any issues I had understanding the online portions were cleared up during the weekly phone call.”*
>
> *“Talking with someone behind a screen is not personal, and it is hard to communicate your true struggles this way.”*
>
> *“Would help to have more of a back and forth with the person”*
>
> *“There are benefits that come from a voice-to-voice conversation that cannot be reproduced by typing into a prompt box.”*

Subtheme: Psychological Challenges

Challenges with participants’ mental health and executive functions such as stress, time management, organization, and problems with memory impacted individuals’ ability to complete the e-CBT program.

> *“Remembering when things needed to get done and setting time aside to do it” “Some, like myself, do not always understand terms or wording on certain subjects” “Difficulty staying on track during stressful life events”*

## Discussion

Although the use of AI in mental healthcare has been supported quantitatively, it is not being integrated to its full potential within the mental healthcare system.^25,26^ As a result, the current study qualitatively examined individuals’ perspectives on the use of an AI-assisted triage system. Furthermore, participants’ experiences within an eCBT program were examined.

Despite there being some support for the use of an AI-assisted triage system, due to previous difficulties in accessing traditional services, many participants expressed concerns with the use of AI for triaging. There was a large emphasis on the lack of ability for AI to recognize the complexities of humans and the need for human oversight to ensure accuracy. Some preferred human triaging since there was a human component that could be more personal and meet the individual needs of people. As a result, providers should check the AI’s recommendations to ensure accuracy during the triage process.

Many clients expressed that they would be fine with the use of AI during the triage process if it allowed them to get expedited access to mental health services. However, they would not be interested in the use of AI throughout psychotherapy. This emphasizes the importance of the therapeutic alliance and human contact within psychotherapy and the client’s desired limitations of its use within the mental health system. Some clients were confused about how AI was being utilized throughout the program, and it was clear that some thought it was used for more than just the triage process. Therefore, education on the use of AI within the triage process could help clients better understand the use of AI as a tool that can expedite their care, in contrast to a replacement for human care. Therefore, the creation of resources to provide clients with a full understanding of the specific use of AI within the triage process could help clients better understand the use of the system.

Similarly, participants had varying experiences with the digital psychotherapy program. One of the main barriers to the program that likely increased participants’ desire to discontinue was technological difficulties. Therefore, further revision of the program to make it more user-friendly and intuitive is important. This could also include implementing more accessible features such as speech-to-text. Another modification of the program could be making it more user-friendly for neurodiverse populations and having modules for autism spectrum disorder and attention deficit hyperactivity disorder. Furthermore, some participants found the program to be repetitive and therefore, psychoeducation could be used at the beginning of the modules to explain the reason for some of the repetition. Overall, people seemed to enjoy their care providers’ feedback since they found it to be helpful and reassuring. However, many participants commented on how increasing human contact would be beneficial since the people who had weekly calls found that this element kept them going throughout the program. However, when implementing more human contact, it is important that it does not only remain behind a screen and that there is voice-to-voice communication. This could include having an introductory call and a final call once the individual has finished all the digital psychotherapy sessions. Furthermore, the digital psychotherapy program currently sends out reminders to clients when their session due date is approaching. However, further helping individuals with executive functioning such as organization and staying on top of their sessions could help allow them to be successful and have a smooth journey within the program.

One of the limitations of this study is selection bias since participants were only recruited from people who previously participated in the corresponding study and were from an outpatient psychiatry waitlist. However, all of these clients actively engaged in the AI system and were capable of commenting on the program’s efficacy. It would be ideal to conduct more studies on the AI system in different populations and use a larger sample. There is also potential for self-selection bias since the survey was optional and it is possible that people who chose to do it might have been the ones who were more open to sharing their experience. Participants also experienced different levels of care within the digital psychotherapy program such as receiving weekly phone calls, receiving only eCBT, or for some, seeing a psychiatrist. This could have impacted individuals’ differing perceptions about the quality of care that they were provided.

Future studies should investigate the best ways to improve communication between caregivers and patients around the use of AI-assisted tools along with the benefits and implications of it. Further research should also be conducted on a larger sample size and within various populations and generations to gather information on differing perspectives about the use of AI within different aspects of the mental healthcare system. Research should also examine the specific levels of care that clients were provided with, to see if this impacts their experiences with the program.

In conclusion, this study examined participants’ perceptions of an AI-assisted triage system and a digital psychotherapy program. Responses were mixed for the use of an AI-assisted triage system and emphasized the importance of human oversight and personalization to ensure accuracy and meet the individual needs of clients. Participants also expressed appreciation for the use of AI in triaging and the ability to access care at an expedited rate. Furthermore, participants expressed various ways that digital psychotherapy programs could be improved including making them more accessible for neurodiversity, improving the user-friendliness of the platform, and doing more psychoeducation on the reasons behind aspects like repetition. They also shared their enjoyment of the ability to receive care as they waited to see a psychiatrist and enjoyed the accessibility of the program.

## Data Availability

Data can be made available upon request.

## Abbreviations

AI: Artificial intelligence
eCBT: Electronic cognitive behavioural therapy
HDH: Hotel Dieu Hospital
KHSC: Kingston Health Sciences Centre
ML: Machine learning
NLP: Natural language processing
OCI: Ontario Centre of Innovation
OPTT: Online Psychotherapy Tool
PI: Principal Investigator
QUOPL: Queen’s University Online Psychotherapy Lab
QI: Quality improvement

## Supplementary Materials

